# Seroprevalence of SARS-COV-2 Antibodies in Scottish Healthcare Workers

**DOI:** 10.1101/2020.10.02.20205641

**Authors:** Hani Abo-Leyah, Stephanie Gallant, Diane Cassidy, Yan Hui Giam, Justin Killick, Beth Marshall, Gordon Hay, Thomas Pembridge, Rachel Strachan, Natalie Gallant, Benjamin J Parcell, Jacob George, Elizabeth Furrie, James D Chalmers

## Abstract

**Introduction:** Healthcare workers are believed to be at increased risk of SARS-CoV-2 infection. The extent of that increased risk compared to the general population and the groups most at risk have not been extensively studied.

**Methods:** A prospective observational study of health and social care workers in NHS Tayside (Scotland, UK) from May to September 2020. The Siemens SARS-CoV-2 total antibody assay was used to establish seroprevalence in this cohort. Patients provided clinical information including demographics and workplace information. Controls, matched for age and sex to the general Tayside population, were studied for comparison.

**Results:** A total of 2062 health and social care workers were recruited for this study. The participants were predominantly female (81.7%) and 95.2% were white. 299 healthcare workers had a positive antibody test (14.5%). 11 out of 231 control sera tested positive (4.8%). Healthcare workers therefore had an increased likelihood of a positive test (odds ratio 3.4 95% CI 1.85-6.16, p<0.0001). Dentists, healthcare assistants and porters were the job roles most likely to test positive. Those working in front-line roles with COVID-19 patients were more likely to test positive (17.4% vs. 13.4%, p=0.02). 97.1% of patients who had previously tested positive for SARS-CoV-2 by RT-PCR had positive antibodies, compared to 11.8% of individuals with a symptomatic illness who had tested negative. Anosmia was the symptom most associated with the presence of detectable antibodies.

**Conclusion:** In this study, healthcare workers were three times more likely to test positive for SARS-CoV-2 than the general population. The seroprevalence data in different populations identified in this study will be useful to protect healthcare staff during future waves of the pandemic.

## Introduction

Healthcare workers (HCWs) are known to be at increased risk of symptomatic infection with severe acute respiratory syndrome coronavirus-2 (SARS-CoV-2)^1,2^. HCWs accounted for 21% of SARS cases during the outbreak in 2002^3^ and high rates of symptomatic infections have been reported across Europe during the present pandemic, including in the UK^4^. Measures taken to mitigate this increased risk include adequate personal protective equipment (PPE)^5^, infection prevention and control (IPC) procedures within healthcare environments and staff testing. Across the UK, testing for healthcare and other key workers with symptoms has been widely available since April 2020^6^.

A key challenge in containing the spread of SARS-CoV-2 has been the potential for asymptomatic or atypical infection^7^. Even in the case of symptomatic individuals reverse transcriptase polymerase chain reaction (RT-PCR) on nasopharyngeal, oropharyngeal or combined upper airway swabs has a reported sensitivity of 70-90% and consequently will underestimate the number of infected individuals^8^. Therefore, the extent of infections in HCWs in different parts of the world remain largely unknown.

Serological testing can be used to determine the incidence and prevalence of SARS-CoV-2 infection^9^. Identifying the extent of healthcare worker infections and the proportion of undetected infections is important to inform IPC measures during future waves of the pandemic.

In this study, we investigated the seroprevalence of SARS-CoV-2 antibodies in a large population of Scottish HCWs.

## Methods

We conducted a prospective observational study recruiting HCWs employed within the National Health Service in Tayside (NHS Tayside). NHS Tayside is a NHS board in the East of Scotland that is responsible for delivering healthcare for over 400,000 people and employs around 14,000 staff.

Healthcare staff were invited to participate in the study via advertisements, including email newsletters and posted adverts on the staff intranet page. Recruitment took place during a single study visit at Ninewells Hospital, which is the health board’s largest teaching hospital. Recruitment took place between 28^th^ May 2020 and the 2^nd^ September 2020. All participants gave written informed consent to participate. The study was approved by the West of Scotland Research Ethics committee, approval number 20/WS/0078

The inclusion criteria were: Employment as a health or social care worker and age over 16 years. Participants were excluded if they had any contraindication to venepuncture, and symptoms consistent with current SARS-CoV-2 infection at the time of enrolment or had tested positive for SARS-CoV-2 in the preceding 14 days.

At the study visit, participants completed a questionnaire on demographics, previous symptoms, employment role, hours of work, contact with patients with COVID-19 infection and whether they had previously tested positive for SARS-CoV-2. Blood samples were taken for measurement of SARS-CoV-2 antibodies in serum.

### SARS-CoV-2 antibody detection

The Siemens SARS-CoV-2 total antibody assay was used in this study. This is a one stop bridging chemiluminescent immunoassay (CLIA) method that detects antibodies against the receptor-binding domain (RBD) of the SARS-CoV-2 spike (S1) protein. The assay is performed on the Siemens Atellica 1300 platform. Validation of this assay was approved by the NHS Scotland national laboratories programme quality group and was then further validated against other commercial antibody platforms in a previous study and found to have 95-100% sensitivity while titres remained constant beyond 81 days following a positive PCR test result^10^.

### Population control subjects

A random selection of blood samples taken at NHS Tayside General Practice Surgeries were tested covering the same time period as the MATCH study cohort. Samples were age and sex matched to the Scottish population demographics to provide a representative sample of the local population to determine the background prevalence of SARS-CoV-2. Serum samples were run on the same Siemens analyser described above.

### Statistical analysis

Data was analysed using IBM SPSS v25 and GraphPad Prism 8.1.2. Chi-squared and Fisher’s test were used as appropriate to compare proportions between groups. Logistic regression was used to derive the odds ratio values for the reported symptom analysis. A p-value <0.05 was considered statistically significant for all analyses.

## Results

A total of 2062 health and social care workers were recruited for this study. The participants were predominantly female (81.7%) and 95.2% were white. The mean age of participants was 44.8 years. Table 1 presents the demographic characteristics of the study participants and their healthcare roles.

**Table 1:**
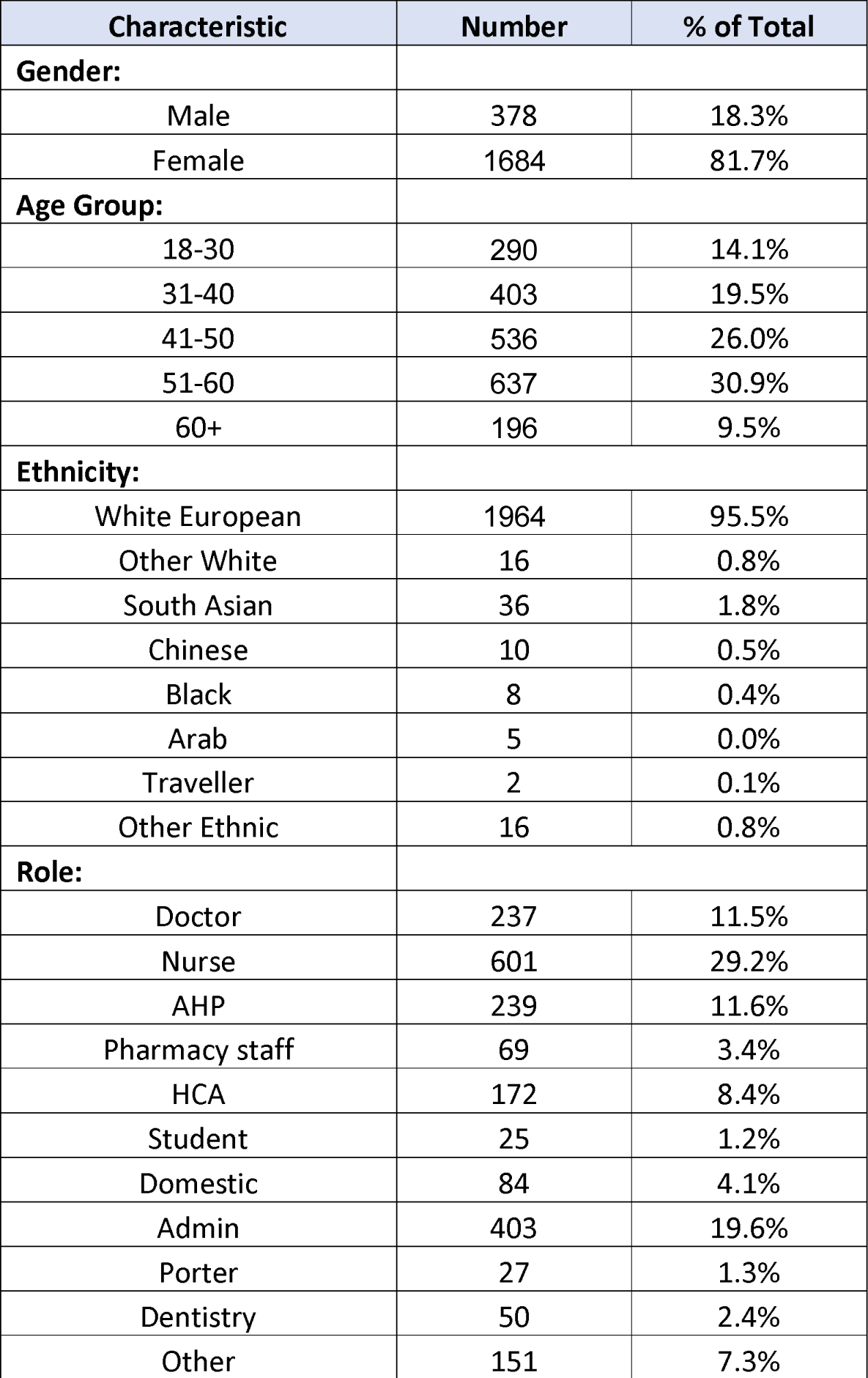
Demographic characteristics of study participants and roles. AHP= Allied health professional, HCA= Healthcare assistant. Other role includes: Lab technician, health scientist, maintenance, laundry, medical physics, other technician, patient transport, chaplaincy, volunteers. Note numbers for ethnicity do not add up to 2062 as participants could chose not to provide this data.

### Seroprevalence of SARS-CoV-2 antibodies

In our study, 299 HCW’s had a positive antibody test directed against SARS-CoV-2 spike protein. This represents a seroprevalence of 14.5%. 11 out of 231 control sera tested positive (4.8%) which was consistent with the broader Scottish surveillance data reported by Health protection Scotland (214 positive tests out of 4751, 4.5%). Compared to both sets of population controls, HCWs had a greater than 3 times greater odds of a positive test (odds ratio 3.4 95% CI 1.85-6.16, p<0.0001 compared to local controls) and (OR 3.6, 95% CI 2.99-4.32, p<0.001 compared to Scotland wide controls).

Table 2 shows the seroprevalence rate amongst subgroups in our study characterised demographic, job role and area of work. Male gender was more frequently associated with detected antibodies (18.5% vs 13.6%, p=0.02). Some job roles were significantly associated with a higher rate of SARS-CoV-2 antibody detection. Healthcare workers in dentistry were the most frequently associated with detected antibodies (26%), followed by Health care assistants (HCA’s) (23.3%) and hospital porters (22.2%), p<0.0001 when comparing across groups. Figure 1 displays the rates of antibody prevalence amongst the HCW by profession.

**Table 2:**
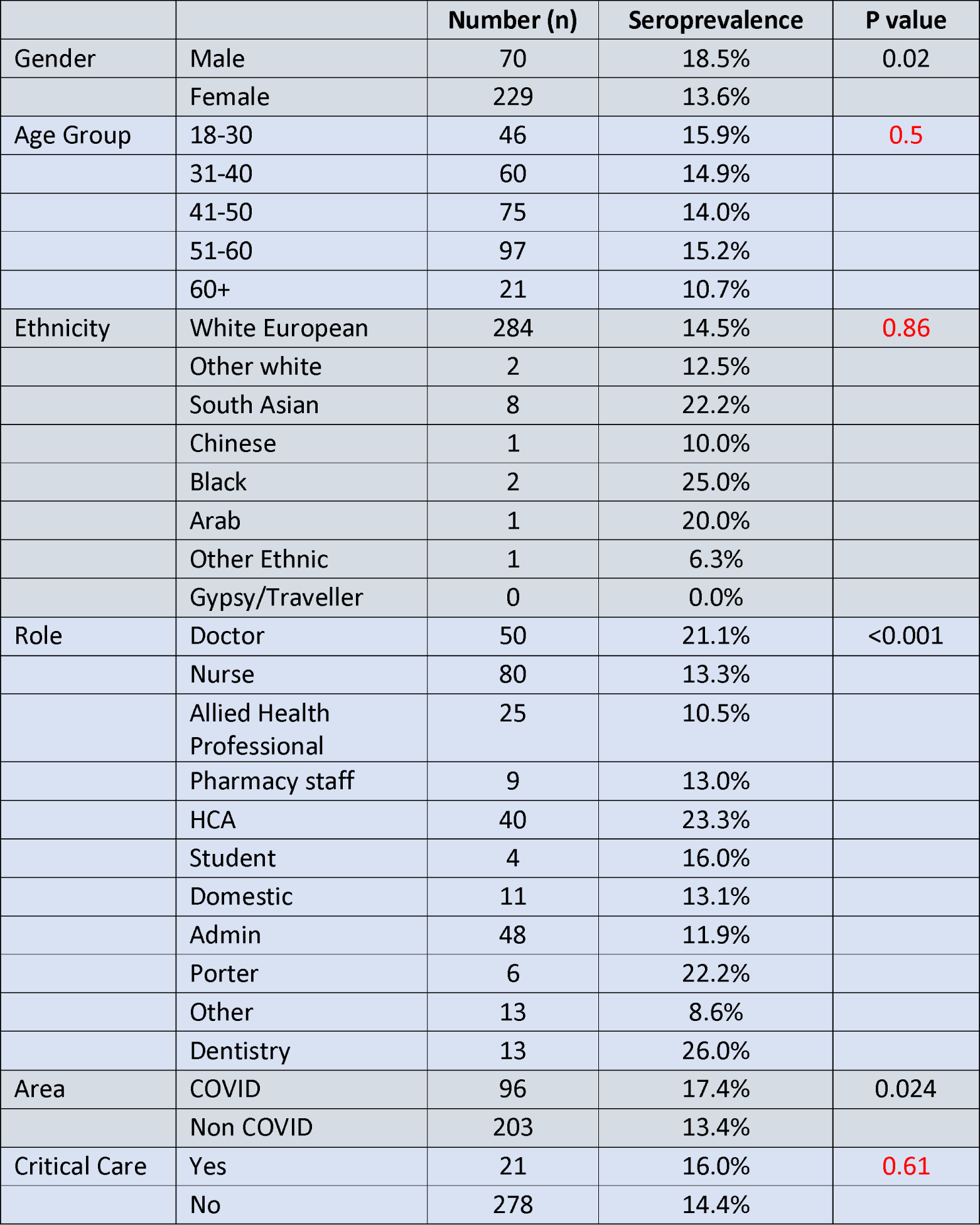
Seroprevalence rate by demographic, role and area of work. Non-significant p values in red font.

**Figure 1:**
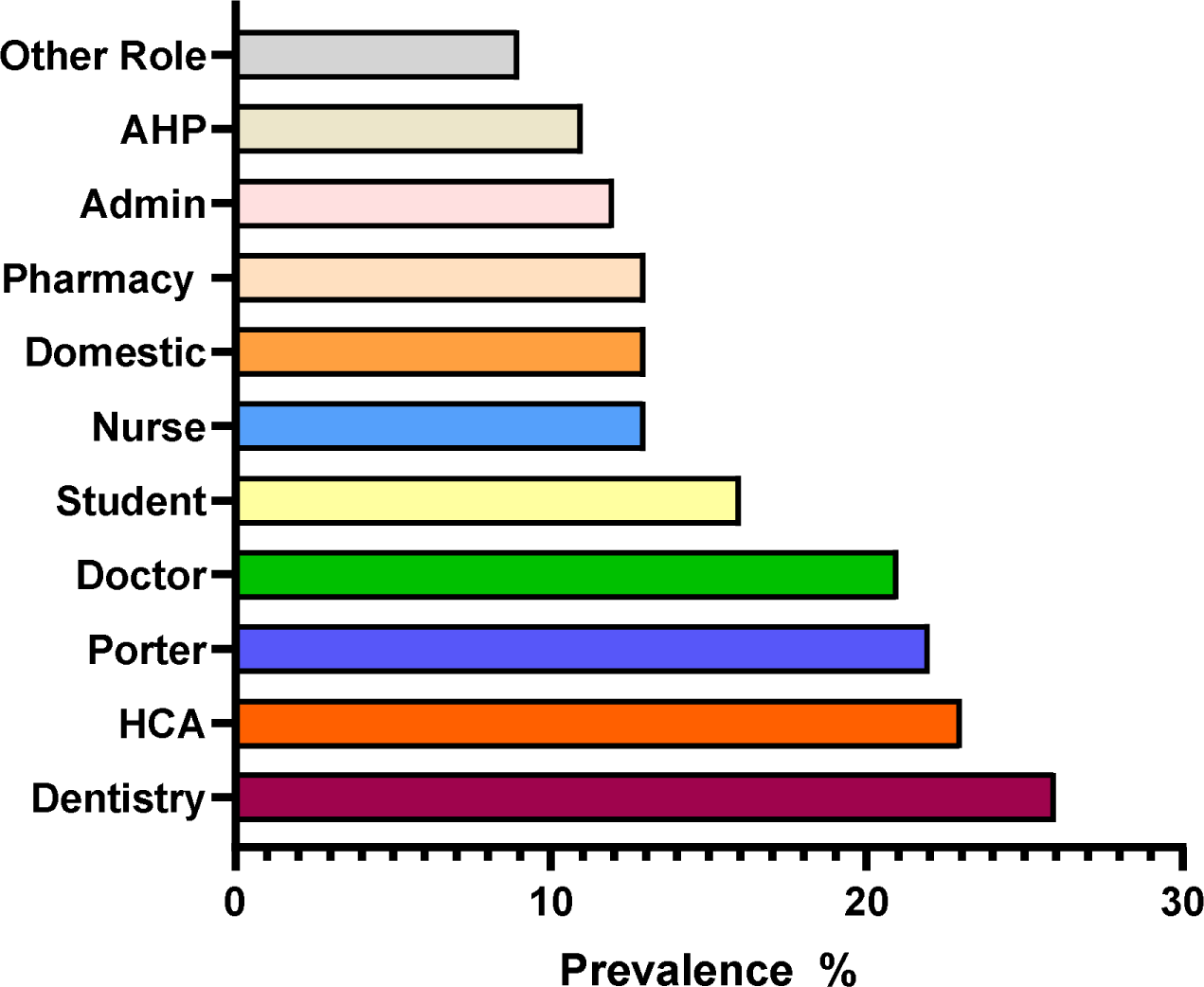
Percentage prevalence of SARS-COV-2 antibody amongst different HCW roles.

Healthcare staff who worked in areas of the hospital that treated suspected or confirmed cases of COVID-19 were more frequently associated with detected antibodies (17.4% vs. 13.4%, p=0.02). Staff who worked in critical care and the intensive care unit were not more frequently associated with detected antibodies (16% vs. 14.4%, p=0.61).

### Prior positive test results and symptomatic infections

Only 624 study participants had ever had a SARS-COV-2 RT-PCR swab. 97.1% (102/105) of participants with positive RT-PCR had detectable antibodies. 11.8% of PCR negative participants had detectable antibody levels. In those who never had a RT-PCR test 9.5% of them had detectable antibodies. Figure 2 displays the proportion of PCR positive and PCR negative participants with detected antibodies.

**Figure 2:**
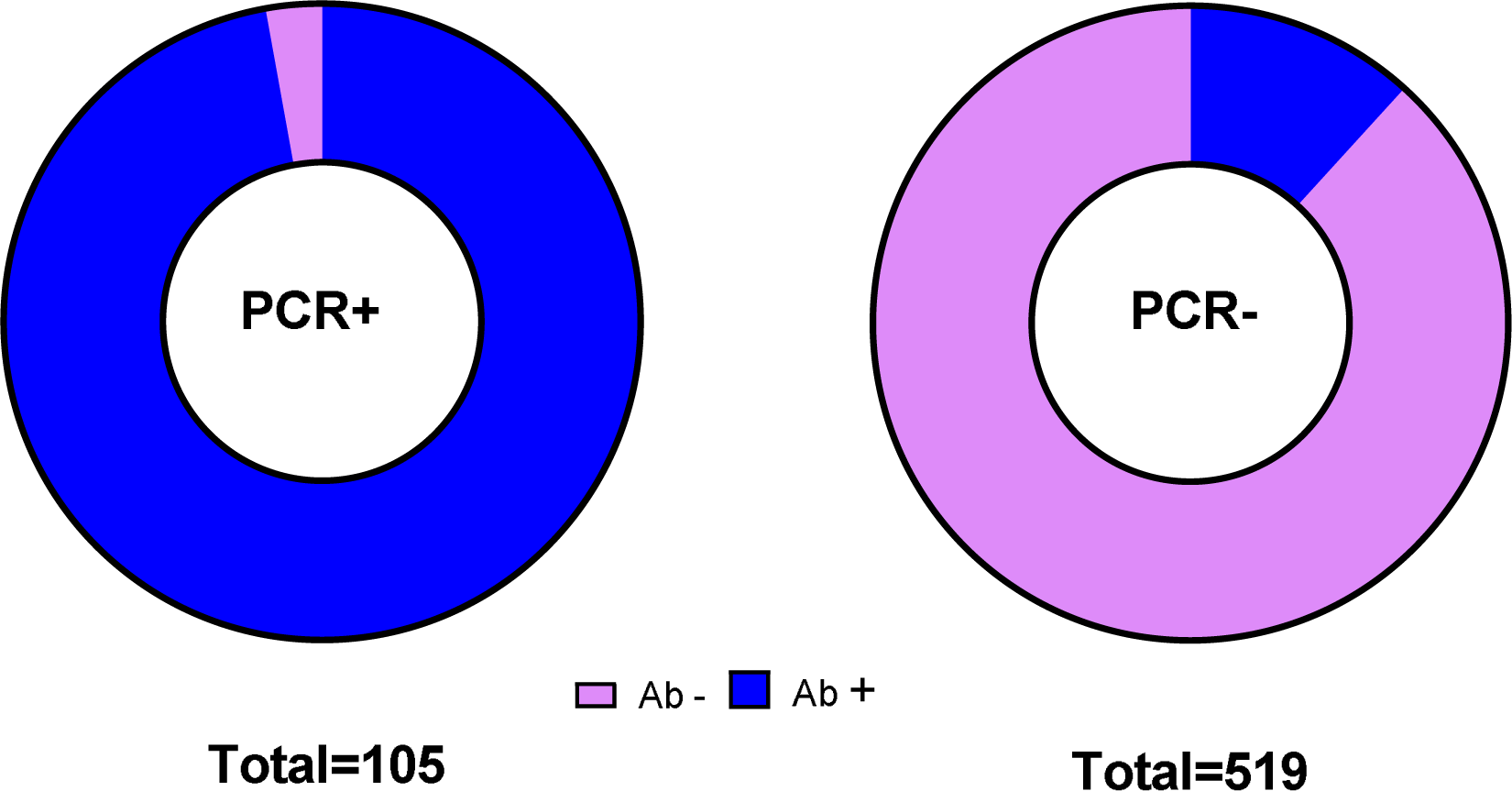
Proportion of antibody detection according to PCR status.

45.4% (n=936) of the HCW’s recruited believed they had COVID-19 but only 25.1% (n=235) of these HCWs had detectable antibodies. Conversely, 18.7% (n=56) of those who had antibodies detected did not believe they ever had COVID-19.

5.1% (n=56) of participants who reported no symptomatic illness during the study period had detectable antibodies compared to 25.6% (n=243) of participants who had at least one symptomatic illness. When compared with the general population individuals who did not have a symptomatic illness during the period of the study did not have an increase frequency of SARS-CoV-2 antibodies (OR 1.20 95% CI 0.30-4.83, p=0.26).

Anosmia was the self-reported symptom that was most likely to correspond with detected antibodies (OR 12.3, 95% CI [9.3-16.3], p <0.001) but was only reported in 5.8% of HCW’s at any time (Table 3). A combination of cough, fever and anosmia was only reported at a frequency of 2.6% but when present was associated with a 10-fold increase in odds of detectable antibody (OR 9.7, 95% CI [6.4-14.7], p <0.001). The absence of any cough, fever and anosmia was associated with a low odds of having a positive antibody test (OR 0.18, 95% CI [0.14-0.24], p <0.001).

**Table 3:**
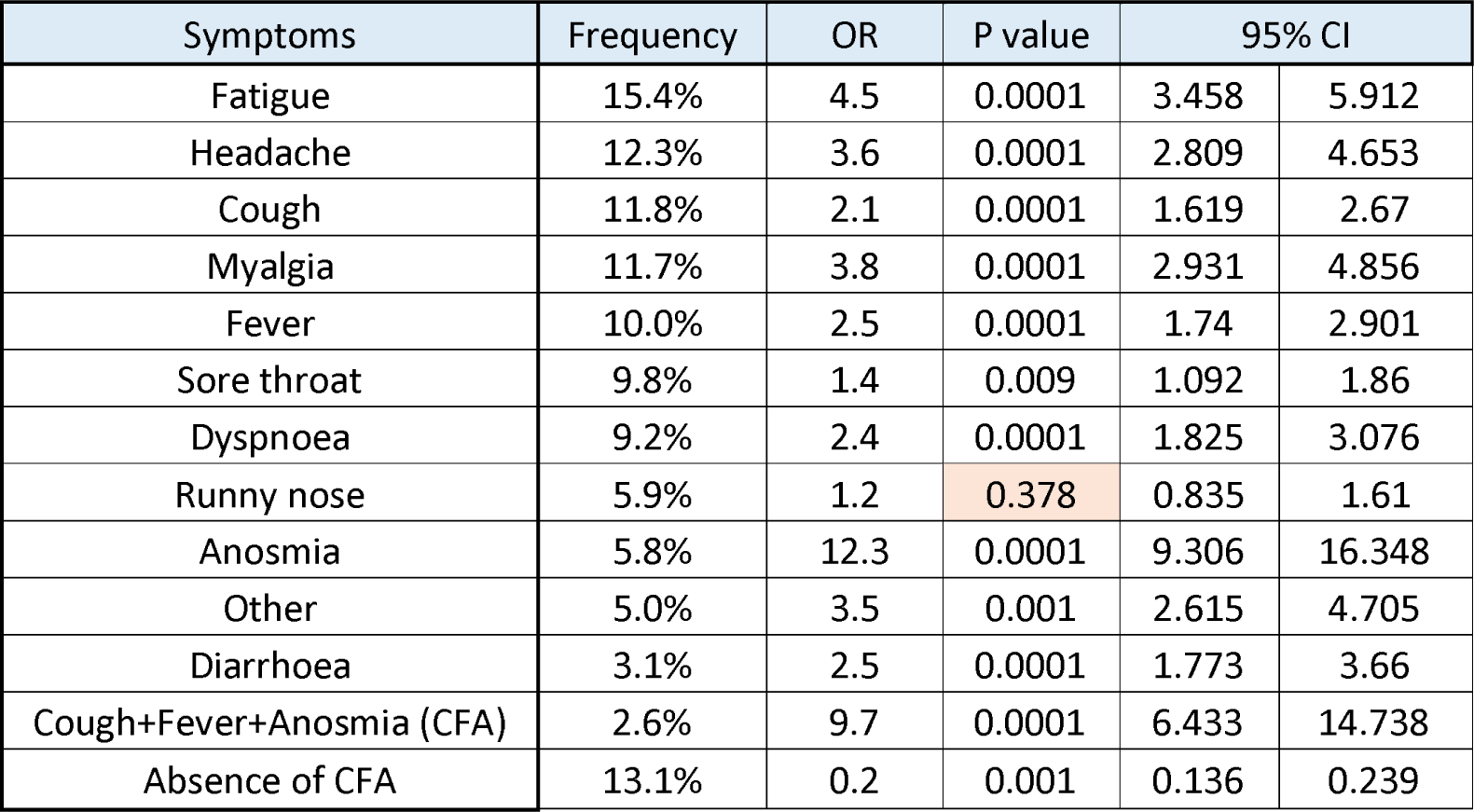
Frequency of reported symptoms and Odds ratio (OR) of each symptom corresponding to a detectable antibody against SARS-COV-2. CFA= combination of cough, fever and anosmia being reported. CI= 95% confidence interval. See appendix for list of other symptoms reports.

| Symptoms | Frequency | OR | P value | 95% CI |  |
| --- | --- | --- | --- | --- | --- |
| Fatigue | 15.4% | 4.5 | 0.0001 | 3.458 | 5.912 |
| Headache | 12.3% | 3.6 | 0.0001 | 2.809 | 4.653 |
| Cough | 11.8% | 2.1 | 0.0001 | 1.619 | 2.67 |
| Myalgia | 11.7% | 3.8 | 0.0001 | 2.931 | 4.856 |
| Fever | 10.0% | 2.5 | 0.0001 | 1.74 | 2.901 |
| Sore throat | 9.8% | 1.4 | 0.009 | 1.092 | 1.86 |
| Dyspnoea | 9.2% | 2.4 | 0.0001 | 1.825 | 3.076 |
| Runny nose | 5.9% | 1.2 | 0.378 | 0.835 | 1.61 |
| Anosmia | 5.8% | 12.3 | 0.0001 | 9.306 | 16.348 |
| Other | 5.0% | 3.5 | 0.001 | 2.615 | 4.705 |
| Diarrhoea | 3.1% | 2.5 | 0.0001 | 1.773 | 3.66 |
| Cough+Fever+Anosmia (CFA) | 2.6% | 9.7 | 0.0001 | 6.433 | 14.738 |
| Absence of CFA | 13.1% | 0.2 | 0.001 | 0.136 | 0.239 |

## Discussion

In this observational study of health and social care workers the SARS-COV-2 seroprevalence rate was 14.5%. To our knowledge this is the first such seroprevalence study to be published in Scotland.

At the time of writing Health Protection Scotland (HPS) is leading a surveillance study for COVID-19 in Scotland on behalf of the Scottish Government^11^. Present data from this study demonstrates a comparative seroprevalence of 4.5% nationally. Our data suggest therefore that health and social care workers are greater than 3 times more likely to be positive for spike protein antibodies and therefore likely to have been infected SARS-CoV-2.

Other HCWs seroprevalence studies conducted in the rest of the UK also reported higher seropositivity when compared to the general population with seroprevalence rates of 24.4%^12^, 25.4%^13^ and 10.7%^14^.

Our study was notable for the inclusion of dentistry staff, who had the highest seroprevalence rate at 26%. This was well above the average seroprevalence rate of 14.5% amongst our HCW’s. Dentistry staff are expected to be a higher risk group given their focus of work is more likely to be aerosol generating and with close exposure to potentially infected mucosal surfaces^15^. Health care assistants, including such staff at nursing homes, had the second highest prevalence at 23%. The caring roles of these workers necessitate close patient contact and their increased risk was also reported in a recent Swedish study^16^.

In other published HCW studies^12^, domestic or housekeeping staff had the highest seroprevalence of antibodies. This was not evident in our study where domestic staff had a below average seroprevalence rate of 13.1%. Other groups who had a seroprevalence rate above average were hospital porters and doctors. Doctors and porters are typically exposed to multiple patients in different working areas. By comparison nurses typically care for up to 6 patients in a defined area during their working day. The variability in work location in a particular time period could be a factor in the increased infection rate we observed.

In our study, HCWs who worked in COVID-19 areas of the hospital had a slightly higher seroprevalence than those workers who did not. This finding is consistent with other studies, including HCW studies conducted in major urban areas where the community burden of COVID-19 was a significant source of exposure^17^. Working in COVID-19 areas of the hospital is one way to define high risk exposure. Nevertheless, the majority of SARS-CoV-2 infections detected in this study occurred in staff who were not working directly with COVID-19 patients, and even so, this group still had a significantly higher prevalence of SARS-CoV-2 antibodies than the general population. While a seroprevalence study cannot establish the source of infection this strongly suggests transmission between healthcare staff within non-clinical environments since many staff roles that did not involve direct contact with patients were still associated with an increased rate of antibody positivity. Therefore, while much media attention has focused on the importance of PPE for front line staff, this data emphasises the importance of IPC measures in non-clinical areas within healthcare environments such as hospitals. The relative success of measures to protect high-risk frontline staff is illustrated by the low rate of antibody detection in critical care staff. All staff working in critical care areas wore PPE in accordance with Health Protection Scotland guidance on working in aerosol generating procedures^18^. We found no significant increased risk of infection for these staff. In the recently published study from Birmingham, UK^12^ staff in intensive care had a significantly lower risk of seropositivity.

We asked our study participants to report if they thought they had COVID19 and list the symptoms they experienced. We describe that only one in four participants who thought they had contracted COVID19 demonstrated serological evidence of infection. The heightened suspicion of infection is justified amongst HCWs, but perceived infection does not correlate well with actual infection. This has some potentially important implications when considering the issue of chronic symptoms in individuals who believe they have had COVID19 infection^19^. However, we demonstrated in this study that certain symptoms are significantly more predictive of infection with SARS-CoV-2. Anosmia represented twelve-fold increased odds with having serologic evidence of infection. This particular symptom was also shown to be strongly predictive in similar analysis performed in other European studies^16,20^.

Our study demonstrates that approximately one fifth (18.7%) of seropositive HCW’s were completely asymptomatic during the study period. This is consistent with studies that only recruited asymptomatic HCW’s^12^ and suggests that a significant proportion of the healthcare workforce will attend work without knowing that they may potentially transmit the infection their colleagues.

This study has potential limitations, including potentially that individuals more likely to believe they have had a SARS-CoV-2 like illness would be more likely to volunteer for such a study. Nevertheless, we were successful in enrolling participants who had never experienced a symptomatic infection and demonstrate an increased seroprevalence even amongst this group. We enrolled patients up to September 2020 and therefore potentially up to 4-5 months post-infection. This raises the possibility of antibodies waning over time^21^. This seems unlikely as a prior study found no evidence of waning of the Siemens assay over 4 months^10^, while a similar Total spike protein antibody assay showed no waning over time in a study from Iceland^22^. Other studies have reported increased infection rate in the BAME population^23,24,25^, we were unable to investigate this as NHS Tayside has a workforce which is 97% white. Important strengths of the study including the large sample sample size, representation of multiple staff groups and the extensive SARS-CoV-2 testing of symptomatic healthcare workers in the region allowing correlation between antibody testing and prior SARS-CoV-2 RT-PCR.^6^

In conclusion our study suggests that HCW are at increased risk of infection with SARS-COV-2 compared with the general population. Our study suggests a differential risk amongst hospital staff and a high proportion of undetected symptomatic and asymptomatic infections. This will help to inform targeted IPC strategies to protect healthcare staff and patients during future waves of the pandemic.

## Data Availability

Data are available from the corresponding author on reasonable request

## Contributors

Study design and conduct: SG, DC, BP, JG, EF, JDC

Data collection: HAL, SG DC, RS, NG, EF, JDC

Laboratory work: DC, YHG, JK, BM, GH, TP, RS, EF

Wrote the manuscript: HAL, JDC

Reviewed the manuscript and approval the final version: all authors

## Notes

Funding: NHS Tayside COVID-19 Research Fund, JDC is supported by the British Lung Foundation Chair of Respiratory Research.

### Competing Interest Statement

JDC reports grants and personal fees from GlaxoSmithKline, Boehringer-Ingelheim, Astrazeneca, Pfizer, Bayer Healthcare, Grifols, Napp, Insmed and Zambon outside the submitted work; All other authors report no conflicts of interest.

### Clinical Trial

not a clinical trial

### Funding Statement

NHS Tayside COVID-19 Research Fund, JDC is supported by the British Lung Foundation Chair of Respiratory Research.

### Author Declarations

The study was approved by the West of Scotland Research Ethics committee, approval number 20/WS/0078

